# Identification of noninvasive and disease-specific biomarkers in hereditary angioedema using urinary proteomics

**DOI:** 10.1101/2023.01.03.23284171

**Authors:** Jianqiang Wu, Xiaoyue Tang, Xue Wang, Peng Liu, Nan Zhou, Zejian Zhang, Yang Cao, Shuyang Zhang, Yuxiang Zhi

## Abstract

**Background:** Hereditary angioedema (HAE) is a rare and potentially life-threatening disease. Noninvasive and disease-specific biomarkers are needed for the early diagnosis and clinical management of HAE.

**Objective:** We sought to apply untargeted proteomics profiling and targeted proteomics validation to identify pathogenic mechanisms and candidate biomarkers of HAE.

**Methods:** Data-independent acquisition (DIA)-based proteomics profiling was performed in urine samples of HAE patients and healthy controls. Bioinformatics analysis was used for functional annotation and pathway enrichment of differentially expressed proteins. Furthermore, promising biomarker candidates were validated in another independent clinical cohort using parallel reaction monitoring (PRM) targeted proteomics quantification.

**Results:** Different urinary proteomics profiles were identified among type 1 HAE, type 2 HAE and healthy controls. A total of 401 differentially expressed proteins were identified between type 1 HAE and healthy controls. Bioinformatics analysis showed that several biological processes and pathways were significantly enriched in HAE, including complement and coagulation cascades, cell adhesion molecules, immune response, proteolysis, and bradykinin catabolic process. Moreover, a promising biomarker panel (C1-INH, KNG1 and EGF) were validated in another independent clinical cohort. The area under the curve (AUC) value of this biomarker panel reached 0.910 for HAE diagnosis (sensitivity: 91.7, specificity: 88.9, *P* <0.001).

**Conclusions:** This study describes the first application of a DIA-PRM workflow to identify noninvasive and disease-specific biomarkers in HAE patients. These findings will contribute to the pathogenesis research and biomarker discovery of HAE.

**Key Messages:**

1. Different urinary proteomics profiles were identified among type 1 HAE, type 2 HAE and healthy controls.
2. Several biological processes and pathways were significantly enriched in HAE, including complement and coagulation cascades, cell adhesion molecules, immune response, proteolysis, and bradykinin catabolic process.
3. A urinary biomarker panel (C1-INH, KNG1, and EGF) could be a promising noninvasive diagnostic tool for HAE.

## 1. Introduction

Hereditary angioedema (HAE) is clinically characterized by recurrent attacks of subcutaneous and submucosal swelling, which involves the face, extremities, trunk, genitalia, upper airways, and/or gastrointestinal tract ^1^. Classic HAE is caused by mutation of the SERPING1 gene, which causes plasma protease C1 inhibitor (C1-INH) deficiency ^2^. HAE with C1-INH deficiency can be subdivided into type 1 (decreased plasma levels of C1-INH) and type 2 (normal levels but dysfunction of C1-INH). Type 1 HAE accounts for the majority of HAE cases in China, and the percentage of type 2 HAE cases in China is < 5%. Other types of HAE due to defects in coagulation factor XII, plasminogen, or angiopoietin-1 have not been reported in China ^3^.

HAE is a rare disease with an estimated prevalence of 1/50,000 and is often not recognized ^4^. As abdominal symptoms are common and important clinical features, HAE is easily misdiagnosed, which can lead to unnecessary surgical procedures ^5,6^. Furthermore, all HAE patients are at risk of a laryngeal attack, and more than 50% have laryngeal attacks during their lifetime. Laryngeal edema-induced asphyxiation without timely treatment is the main cause of death in HAE patients, and the mortality rate ranges between 11% and 40% ^7-9^. However, previous studies have reported that the interval between the onset of symptoms and final diagnosis of HAE remains 8-13 years ^4,10,11^. Long diagnostic delay and inappropriate treatment will aggravate the humanistic and economic burden for HAE patients ^12^. Therefore, early diagnosis and disease evaluation are essential to improve the quality of life and clinical management of HAE patients.

Biomarkers are measurable changes associated with physiological or pathophysiological conditions ^13,14^. Without homeostatic control, urine accumulates systematic changes in the body and has the potential to reflect small and early pathological conditions ^14^. Moreover, urine can be collected noninvasively and in large volume. Thus, urine is an attractive sample source for biomarker research. Proteomics is a powerful tool in biomarker discovery, exploration of pathogenesis, and drug target identification ^15^. With the rapid development of mass spectrometry (MS) techniques, urinary proteomics has become a promising field in biomarker discovery ^16^. The urinary proteome is mainly composed of plasma proteins that pass the glomerular barrier and proteins shed by cells within the urogenital system. Thus, urinary proteomics could reflect both systemic and local conditions of the body. Currently, in addition to its application in urological diseases ^17^, urinary proteomics has also been applied in cardiovascular diseases ^18^, brain diseases ^19^ and other diseases ^20-22^. Although urinary biomarkers have been investigated in many diseases, no urine biomarkers are currently available for the diagnosis, disease monitoring and therapeutic efficacy evaluation of HAE ^23^. Advances in high-throughput proteomics technology may facilitate the discovery of promising noninvasive urine biomarkers of HAE.

In this study, the data-independent acquisition (DIA)-based proteomics technique was applied to explore the urinary protein profiling of HAE patients. Bioinformatics analysis was performed to annotate the biofunctions and pathways of the differentially expressed proteins (DEPs). Then, several promising biomarkers were further validated in another independent clinical cohort using parallel reaction monitoring (PRM)-based targeted proteomics. This study will provide a novel noninvasive method for HAE diagnosis and will improve our understanding of the pathogenesis of HAE.

## 2. Methods

### 2.1 Patients and sample collection

In the biomarker discovery phase, 20 adult HAE patients and 29 healthy controls (HCs) were recruited from Peking Union Medical College Hospital (PUMCH) during 2020. Among these patients, 15 had type 1 HAE, and 5 had type 2 HAE. In the biomarker validation phase, 24 other type 1 HAE patients and 18 HCs were recruited from PUMCH during 2021. This study was conducted in accordance with the Declaration of Helsinki (as revised in 2013). All HAE patients and HCs provided written informed consent, and the Ethics Committee of PUMCH approved this study (No. HS-2402).

Midstream first morning urine was collected from these subjects. Routine urine test results were normal in all subjects. After collection, the urine samples were centrifuged at 3,000 × g for 20 min at 4 °C to remove the cell debris, and the supernatants were stored at −80 °C for further proteomics analysis.

### 2.2 Sample preparation

Samples were prepared using a filter-aided sample preparation method as previously described ^24^. Briefly, samples were reduced with 20 mM dithiothreitol at 100 °C for 10 min and alkylated with 50 mM iodoacetamide at room temperature for 30 min in the dark. Then, urinary proteins were extracted from the samples by acetone precipitation, and the pellets were resuspended with 20 mM Tris buffer. Then, 100 μg of protein was loaded onto 30-kD filter devices (Pall, Port Washington, NY, USA) and centrifuged at 14,000 × g at 18 °C. After washing three times with 20 mM Tris buffer, the samples were digested with trypsin at 37 °C overnight. Finally, 2 μg of each peptide sample was used for the subsequent LC□MS/MS analysis.

### 2.3 DIA quantitative proteomic analysis

LC⍰MS/MS analysis was performed by an Orbitrap QE HF mass spectrometer coupled with an EASY-nLC 1200 UHPLC system (Thermo Fisher Scientific, Waltham, MA, USA). Digested peptides were loaded onto a trap column (75 μm × 2 cm, 3 μm, C18, 100 A°). The peptides were separated on an analytical column (75 μm × 500 mm, Kyoto Monotech, Kyoto, Japan). The elution was set to 5–30% buffer B (0.1% formic acid in 99.9% acetonitrile; flow rate of 0.3 μl/min) for 60 min. Moreover, an iRT kit (Biognosys, Schlieren, Switzerland) was added to all samples to make retention time alignments among samples ^25^. The MS was set in DIA mode, and the parameters were set as follows: the full scan was performed with a resolution of 120,000 and in the range of 350–1,500 m/z; the cycle time was 3 s; the automatic gain control (AGC) was 3e6, and the injection time was under 100 ms; precursors were selected by charge state screening with a +2 to + 6 charge state; and the dynamic exclusion duration was 10 s. During the DIA analysis, a mixed sample was inserted after every ten samples for quality control (QC).

### 2.4 PRM proteomics validation

First, pooled peptide samples were subjected to LC□MS/MS analysis with six technical replicates. Skyline software was used to build the spectrum library and screen peptides for PRM analysis. For each targeted protein, two to five peptides were selected using the following criteria: identified with q value < 1%, completely digested by trypsin, 8-18 amino acid residues, exclusion of the first 25□N-terminal amino acids, and carbamidomethylation of cysteine as the fixed modification. Only unique peptides of each protein were used for further PRM quantitation. The retention time (RT) segment was set to ± 2□min for each targeted peptide, with its expected RT in the center based on the pooled sample analysis.

To ensure data quality, a mixed sample was used as the quality control after every 10 samples to determine the stability of the instrument signal. To further ensure data quality, an iRT standard was added to each sample, and the stability of the chromatographic retention time was evaluated during the analysis. To reduce any system bias, different groups of samples were analyzed in a random order.

### 2.5 MS data processing and analysis

All raw MS data in the DIA experiment were imported into Spectronaut software (Biognosys AG, Switzerland) to generate the spectra library. The data were searched against the UniProt human database appended with the iRT fusion protein sequence. The parent ion tolerance was set at 10 ppm, and the fragment ion mass tolerance was set to 0.05 Da. A maximum of two missed cleavage sites in the trypsin digestion was allowed. Carbamidomethylation of cysteines was set as a fixed modification, and the oxidation of methionine was considered a variable modification. Then, the DIA raw files were searched against the self-built spectra library using Spectronaut Pulsar X. The optimal XIC extraction window was determined according to the iRT calibration strategy. The cross-run normalization was set to the local normalization based on the local regression. Protein inference was performed using the implemented IDPicker algorithm ^26^. The Q value was set to 0.01 for data filtering. The sum peak areas of the respective fragment ions in MS_2_ were used to quantify peptide intensities.

For the PRM validation, the MS data were processed with Skyline software. The transition settings in Skyline were as follows: precursor charges, + 2 to + 5; ion types, b, y, and p; the product ions from ion 3 to last ion − 1; ion match tolerance, 0.02□m/z; the six most intense product ions were picked; and the variable “min dotp” was set to 0.7. The correct peaks were selected manually, and all of the peptide results were exported. Total ion current (TIC) chromatograms were obtained for each sample in the range of a +2 to +5 charge using Progenesis QI for Proteomics software. The mass spectra data for each peptide were normalized with the TIC chromatogram of its associated sample to correct for sample size and mass signal intensity errors. The results for each peptide were quantitatively analyzed, and the identified differential proteins were screened and compared with the DIA results.

### 2.6 Bioinformatics and statistical analysis

Functional annotation of differentially expressed proteins (DEPs) was performed using DAVID database ^27^. The biological process, cellular component and molecular function of DEPs were enriched. For Ingenuity Pathway Analysis (IPA), the SwissProt accession numbers were uploaded to IPA software (Ingenuity Systems, CA, United States). The biofunction and canonical pathway of DEPs were enriched. Pattern recognition analysis (principal component analysis, PCA; orthogonal partial least squares discriminant analysis, OPLS-DA) was conducted by SIMCA software (version 15.0, Umetrics, Sweden). The statistical analysis was performed with GraphPad Prism (version 8.0.2, GraphPad Software Inc., San Diego, California, USA). Receiver operating characteristic (ROC) analysis was performed with the “Biomarker discovery” module on the MetaAnalyst 5.0 website (https://www.metaboanalyst.ca/).

## 3. Results

### 3.1 Clinical characteristics of subjects and study design

A total of 44 HAE patients and 47 healthy controls were recruited in this study. In the discovery phase, 20 newly diagnosed HAE patients (type 1: 15 patients; type 2: 5 patients) and 29 HCs were included for differential proteome discovery. HAE patients and healthy controls were matched with age and sex. The average concentration of plasma C1 inhibitors in type 1 HAE patients was significantly lower than that in type 2 HAE patients and HCs. In the validation stage, another independent clinical cohort was recruited for biomarker validation, including 24 type 1 HAE patients and 18 HCs.

### 3.2 DIA-based quantitative proteomics analysis

In the biomarker discovery stage, urinary samples of 20 HAE patients and 29 HCs were analyzed using the DIA quantitative proteomics method. A quality control sample was used to assess the stability of the results. The correlations of the quantified protein intensities of the QC samples showed satisfactory reproducibility (R^2^-values: > 0.90). A total of 2562 protein groups were identified in the urine samples, and 1716 reliable proteins quantified with at least two unique peptides were used for further analysis.

This study compared the urine proteome profiles of HAE patients and HCs. The results showed that there were significantly different urinary proteome profiles among type 1 HAE, type 2 HAE and HCs **(Figure 1A)**. Thus, type 2 HAE may involve different pathophysiological processes compared with type 1 HAE. Interestingly, urinary C1-INH expression was significantly higher in type 2 patients than in type 1 HAE patients (ratio = 2.76, p < 0.001). This result was consistent with the higher expression of plasma C1-INH in type 2 HAE than in type 1 HAE. Moreover, the OPLS-DA results showed that type 1 and type 2 HAE had significantly different proteomic expression in urine samples, and these two disease subtypes could be clearly separated from each other in the model **(Figure 1B)**.

**Figure 1.**
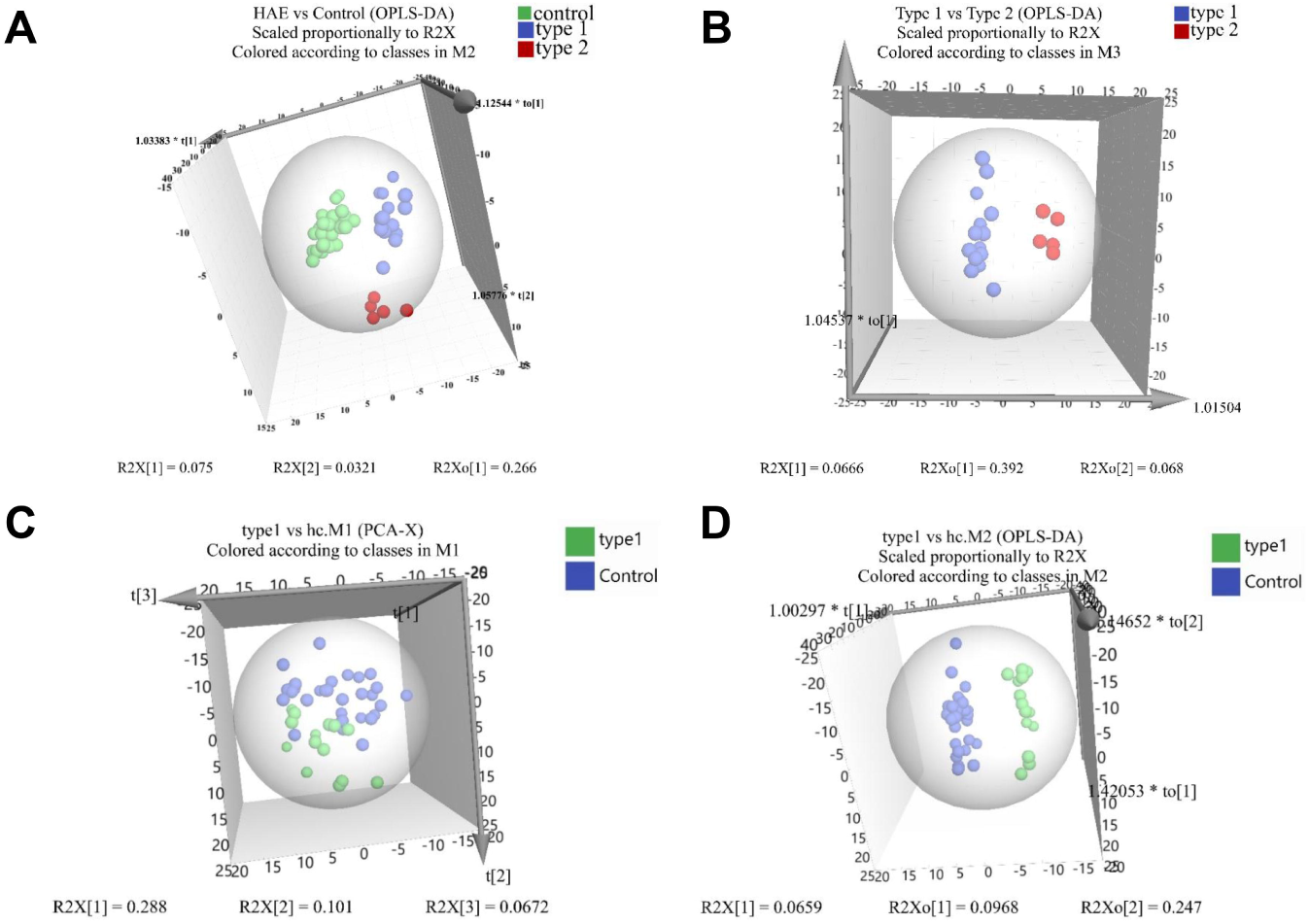
Urinary proteome profiling of type 1 HAE, type 2 HAE and HCs. (A) OPLS-DA of type 1 HAE, type 2 HAE, and heathy controls; (B) OPLS-DA of type 1 HAE and type 2 HAE; (C-D) PCA and OPLS-DA of type 1 HAE and HCs.

Because type 1 HAE accounts for more than 95% of all HAE cases in China, the comparison of the urinary proteome between type 1 HAE and HCs was the focus of this study. The PCA and OPLS-DA results showed that type 1 HAE can be distinguished well from HCs based on the urinary protein pattern **(Figure 1 C-D)**. A total of 401 differential urinary proteins were identified (ratio >1.5, p < 0.05). Interestingly, it was found that the abundance of urinary C1IN in type 1 HAE was significantly lower than that of HCs (Ratio=2.79, p<0.0001), suggesting that urinary C1-INH could be a promising noninvasive diagnostic biomarker of type 1 HAE.

### 3.3 Bioinformatics analysis of DEPs between type 1 HAE patients and HCs

Functional annotation of the differential proteins between type 1 HAE patients and HCs was performed using the DAVID database. In the biological process category, cell adhesion, immune response, proteolysis, complement activation, blood coagulation, innate immune response in mucosa, and bradykinin catabolic process were significantly enriched **(Figure 2A)**. In the molecular function category, antigen binding, calcium ion binding, collagen binding, integrin binding, cadherin binding, extracellular matrix structural constituent, signaling receptor activity, serine-type endopeptidase inhibitor activity, and protease binding were significantly enriched **(Figure 2B)**.

**Figure 2.**
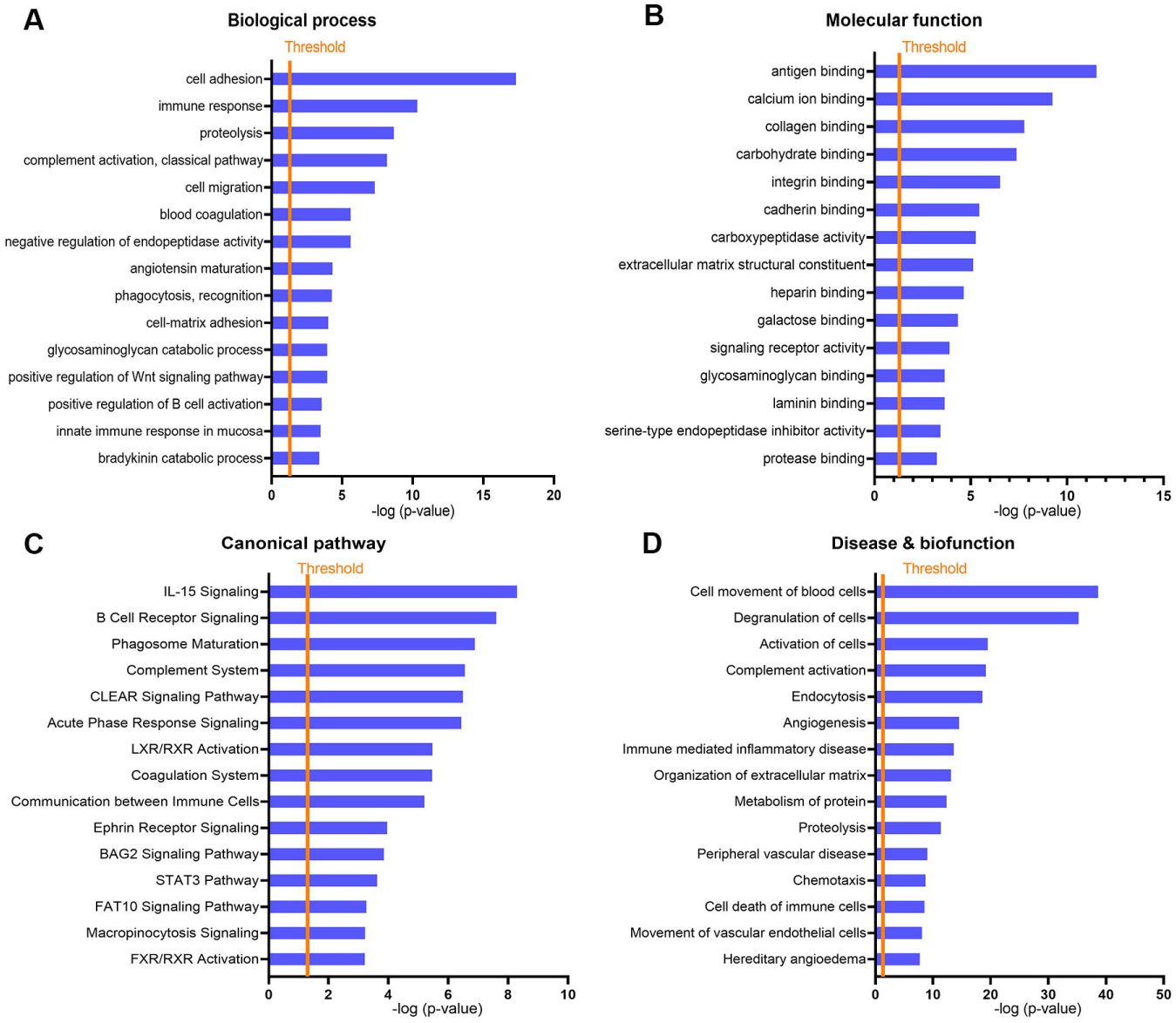
Bioinformatics analysis of DEPs between type 1 HAE patients and HCs. (A-B) Biological processes and molecular functions annotated in the DAVID database; (C-D) canonical pathway and disease & biofunction annotation of DEPs in IPA.

To identify the major canonical pathways and diseases and biofunctions involved with the differential urine proteins, IPA was used for enrichment analysis. It was demonstrated that IL-15 signaling, B-cell receptor signaling, phagosome maturation, complement system, acute phase response signaling, LXR/RXR activation, coagulation system, and communication between immune cells were significantly enriched **(Figure 2C)**. Furthermore, in the disease and biofunction category, cell movement of blood cells, degranulation of cells, activation of cells, complement activation, angiogenesis, immune-mediated inflammatory disease, peripheral vascular disease, movement of endothelial cells, and hereditary angioedema were significantly enriched during HAE pathogenesis **(Figure 2D)**. Interestingly, eight differential proteins identified in our study were annotated in IPA to the disease item “hereditary angioedema”, including angiotensin-converting enzyme (ACE), complement C1s subcomponent (C1S), neutrophil elastase (ELANE), coagulation factor XII (F12), kininogen-1 (KNG1), plasminogen (PLG), alpha-1-antitrypsin (SERPINA1), and plasma protease C1 inhibitor (SERPING1 or C1IN). These results indicated that urinary proteomics reflects the pathophysiological changes that occur in HAE.

### 3.4 Association between the urinary proteome and clinical features

These DEPs were then mapped into a coexpression network using STRING and Cytoscape. The most significant cluster of the DEP coexpression network detected by MCC (Cytoscape, CytoHubba) consisted of 10 hub proteins (CLU, C1-INH, CD59, KNG1, C4A, EGF, RAC1, ACE, ANPEP and ENPEP) (**Figure 3A**). We also analyzed the functions and networks of these hub proteins through GeneMANIA, and the main functions of hub proteins were complement activation and coagulation (**Figure 3B**). In addition, we selected several hub proteins to construct a biomarker combination, and a biomarker panel consisting of plasma protease C1 inhibitor (SERPING1 or C1-INH), pro-epidermal growth factor (EGF) and kininogen-1 (KNG1) had an ideal diagnostic performance with an AUC of 0.956 (sensitivity: 100%, specificity: 82.8%) (**Figure 4A**).

**Figure 3.**
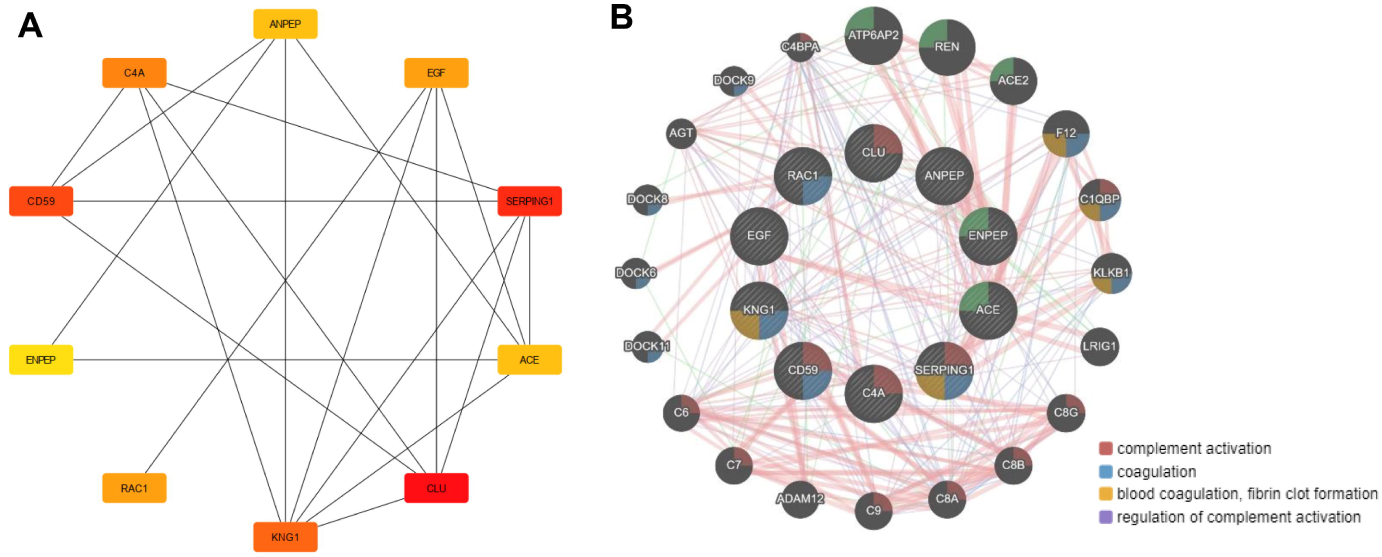
Coexpression network analysis of potential biomarkers of HAE. (A) Ten hub proteins of the coexpression network; (B) the major functions of the hub proteins.

**Figure 4.**
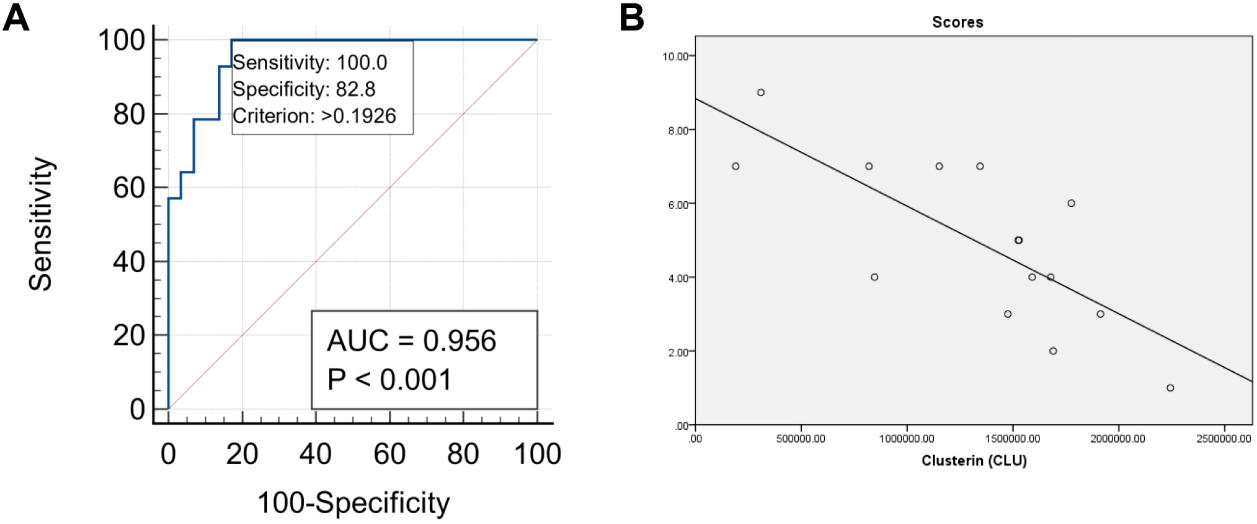
Diagnostic performance and clinical association of candidate biomarkers. (A) The ROC analysis result of a biomarker panel (EGF+C1-INH+ KNG1) in DIA; (B) correlation of the urinary CLU level and disease severity score of HAE.

To demonstrate the clinical utility of our in-depth urinary proteomics analysis, we further analyzed the association between the urinary proteome and disease severity. As previously described ^28^, a severity scoring system was used to assess the disease severity of HAE, which was mainly based on the onset age, edema location, clinical manifestation, and long-term prophylaxis. Our results indicated that the expression levels of two urine proteins (RAC1 and JAM3: junctional adhesion molecule C) were positively correlated with the disease severity score, and the expression levels of six urine proteins (QPCT, ANPEP, B4GALT1, MAN1A1, CLU, and EGF) were negatively correlated with the disease severity score. In particular, CLU was the most significant protein correlated with disease severity (R=-0.758, p=0.001), suggesting that urinary clusterin has the potential to predict the disease severity of HAE (**Figure 4B**).

### 3.5 PRM validation of promising biomarkers of HAE

The biomarker panel was further validated in another independent cohort including C1-INH, KNG1, and EGF. To ensure system stability and limit systemic bias during targeted proteomic analysis, QC samples were injected before and after every 10 samples. By performing PCA on both the QC samples and individual urine samples, the QC samples were closely clustered. The correlation map of QC samples also showed that the QC samples were highly positively correlated with a Pearson correlation coefficient above 0.95. These results demonstrated the stability of the MS platform during proteomics analysis.

According to PRM targeted proteomics quantification, these three proteins showed the same trend as those quantified in the DIA experiment and were significantly decreased in type 1 HAE compared with HCs. Furthermore, the diagnostic performance of this biomarker panel was evaluated using ROC analysis. The results showed that this biomarker panel had good diagnostic performance with an AUC value of 0.910 (sensitivity: 91.7%, specificity: 88.9%) **(Figure 5)**.

**Figure 5.**
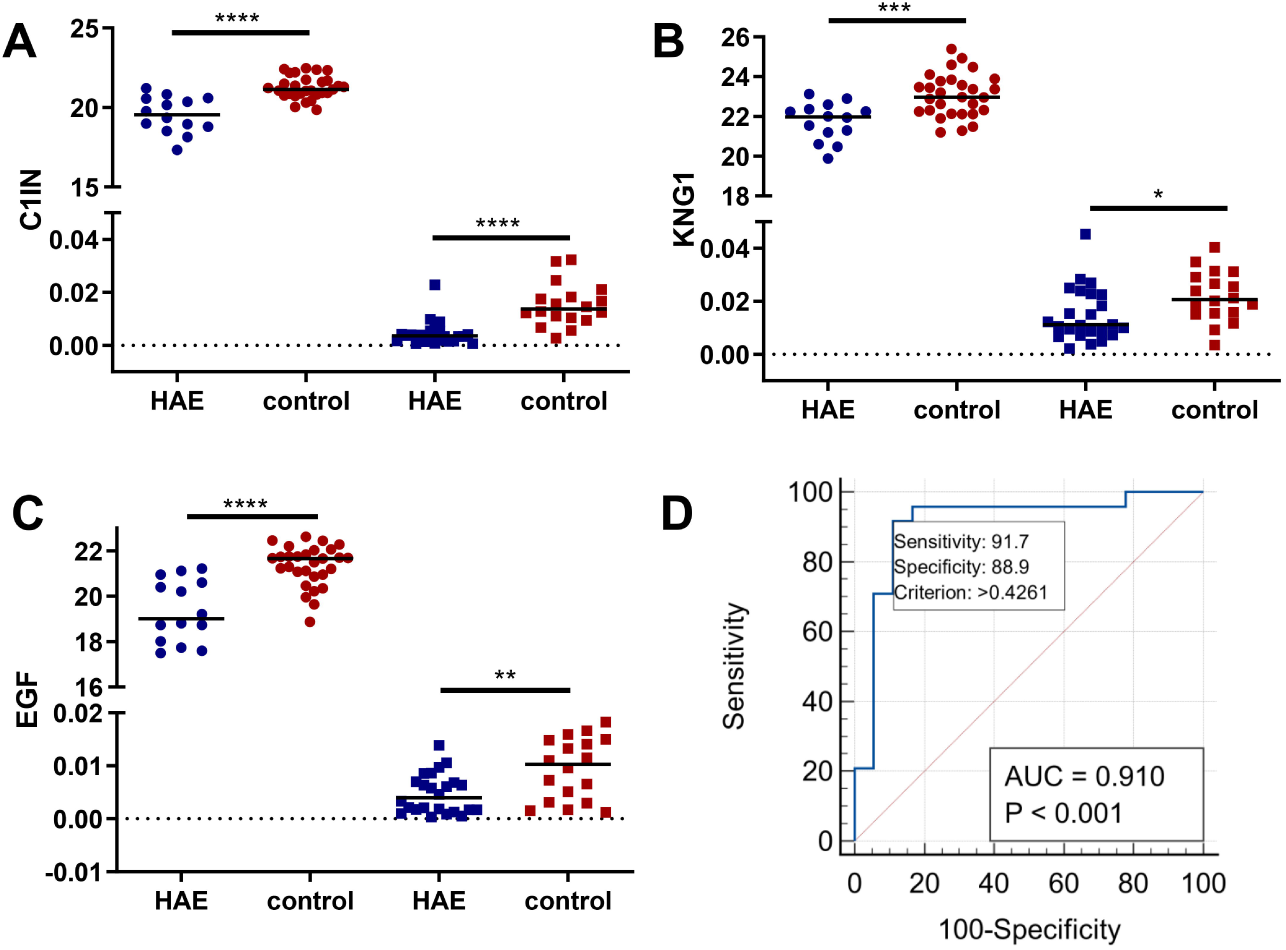
Targeted validation of promising biomarkers in another cohort. (A-C) DIA and PRM proteome quantification of C1IN, KNG1 and EGF in urine samples; (D) ROC analysis of the biomarker panel (C1IN+ KNG1+EGF) in PRM.

## 4. Discussion

In this study, we performed DIA-based urinary proteomics profiling in HAE patients. Several important biological processes and pathways were significantly enriched in the pathogenesis of HAE. Furthermore, some disease-related biomarkers of HAE were identified and further validated in another clinical cohort using PRM-based targeted proteomics quantification. To our knowledge, this is the first application of urinary proteomics analysis in HAE patients, and this study will provide a noninvasive method for HAE diagnosis and improve the understanding of the pathogenesis of HAE.

By DIA proteome profiling, it was found that there were significantly different urinary proteome profiles among type 1 HAE patients, type 2 HAE patients and HCs. Thus, although type 1 HAE and type 2 HAE share the deficiency of C1-INH, type 2 HAE may involve different pathophysiological processes compared with type 1 HAE. As the sample size of type 2 HAE patients is relatively small, the conclusion needs further confirmation. Interestingly, urinary C1-INH expression was significantly higher in type 2 HAE patients than in type 1 HAE patients (ratio = 2.76, p < 0.001), and urinary C1-INH in type 1 HAE patients was significantly lower than that in HCs (ratio=2.79, p<0.0001). The results suggested that urinary C1-INH is a promising noninvasive and disease-specific biomarker for HAE.

HAE disease is characterized by recurrent episodes of mucosal edema. Currently, the pathogenesis of HAE is not well understood. Previous studies have revealed that classic HAE is caused by a deficiency of C1-INH, which can regulate proteases in several pathways, including the complement, contact system, coagulation, and fibrinolytic pathways. Moreover, deficiency of C1-INH and activation of the contact system can lead to bradykinin overproduction. Bradykinin is the central mediator of swelling in HAE, and activation of its receptor on vascular endothelial cells leads to vascular instability, hyperpermeability, plasma extravasation, and resultant angioedema ^29,30^. As type 1 HAE accounts for more than 95% of all HAE cases in China ^11^, functional annotation of differential proteins between type 1 HAE patients and HCs was the focus of this study. Our results showed that several biologic processes were significantly changed in HAE, such as proteolysis, complement activation, blood coagulation, innate immune response in mucosa, and bradykinin catabolic process. Furthermore, complement activation, angiogenesis, immune-mediated inflammatory disease, peripheral vascular disease, movement of endothelial cells, and hereditary angioedema were significantly enriched in the disease and biofunction category. These bioinformatics results were closely related to the pathophysiology of HAE and will lead to a better understanding of the pathogenesis of HAE. Moreover, eight differential urinary proteins identified in our study were annotated in IPA to the disease item “hereditary angioedema”. These results indicated that urinary proteomics reflects the pathophysiological changes that occur in HAE and may serve as a noninvasive biomarker source of HAE.

Because the sensitivity and specificity of a single biomarker are often inadequate, the establishment of a biomarker panel is preferred. In this study, we selected several hub proteins in the coexpression analysis to construct a biomarker panel for HAE diagnosis. In the biomarker discovery cohort, a combination of three urinary proteins (C1-INH + KNG1 + EGF) showed good performance for the diagnosis of HAE with an AUC of 0.956. Moreover, this biomarker panel was successfully validated in another independent clinical cohort with an AUC of 0.910, suggesting its good clinical application prospects. C1-INH has serine-type endopeptidase inhibitor activity and plays a crucial role in regulating important physiological pathways, including complement activation, blood coagulation, fibrinolysis, innate immune response and the generation of kinins. Currently, plasma C1-INH is a crucial disease biomarker of HAE in clinical practice. Furthermore, kininogen is an inhibitor of thiol protease. HMW-kininogen plays an important role in blood coagulation, and bradykinin is released from HMW-kininogen as a result of activation of the contact system ^31^. A recent study reported that mutations in the KNG1 gene can cause a novel type of HAE ^32^. In addition, EGF has rarely been reported in HAE, but EGF can regulate endothelial cell migration and proliferation. Thus, EGF may affect vascular endothelial cell organization and plasma extravasation. The role of EGF in the pathogenesis of HAE requires further research. In addition, we observed that urinary CLU was significantly correlated with the disease severity of HAE (R=-0.758, p=0.001). CLU is a regulator of the complement terminal pathway and can sequester terminal complement components C7-C9 to block the assembly of the membrane attack complex (MAC) ^33^. Thus, urinary CLU is a potential biomarker to predict the prognosis of HAE.

HAE is a rare and potentially life-threatening disease. This disease is often misdiagnosed for years owing to the general lack of knowledge and its nonspecific manifestations. This condition leads to HAE patients not only suffering from the effects of the disease itself but also from ineffective treatment and needless invasive diagnostic and therapeutic medical procedures. To minimize the suffering and improve quality of life, it is necessary to discover noninvasive and specific biomarkers for HAE. In this study, using DIA- and PRM-based proteomics quantification, we identified several promising biomarkers for HAE. Moreover, significant biological processes and pathways were identified in this study, which would provide references for further investigations on the pathogenesis of HAE. As HAE is a rare disease, the limitation of this study is that it was performed at a single center with a relatively small sample size. In the future, multicenter and prospective clinical validation is needed to verify the clinical application of these biomarkers.

## 5. Conclusion

To our knowledge, this is the first application of urinary proteomics in HAE patients. Our results showed that the urinary proteome can reflect the pathophysiological changes of HAE and has the potential to serve as a noninvasive biomarker source for the diagnosis and clinical management of HAE. A biomarker panel (C1HN, KNG1, and EGF) showed good performance for the diagnosis of HAE. This study provides a novel noninvasive method for HAE diagnosis and enhances our understanding of the pathogenesis of HAE.

## Data Availability

All data produced in the present study are available upon reasonable request to the authors

## Abbreviations Used

HAE: hereditary angioedema
HC: heathy control
LC-MS/MS: liquid chromatography coupled with tandem mass spectrometry
DIA: data-independent acquisition
PRM: parallel reaction monitoring
DEP: differentially expressed protein
C1-INH or SERPING1: Protease
C1: inhibitor
KNG1: Kininogen-1
EGF: Pro-epidermal growth factor
CLU: Clusterin
ROC: receiver operator characteristics
AUC: area under the curve

## Acknowledgements

This study was supported by grants from the National Key Research and Development Program of China (No. 2016YFC0901501), the National Natural Science Foundation of China (No. 81472870), the CAMS Innovation Fund for Medical Sciences (CIFMS 2021-1-I2M-003) and Youth Science Fund of Peking Union Medical College Hospital (pumch201912152). The funding organization was not involved in the collection, management, or analysis of the data; preparation or approval of the manuscript; or decision to submit the manuscript for publication.

## Author Contributions

JW, YZ and SZ: proposed and designed this study. JW, XT, XW, PL, NZ, ZZ and YC: collected the samples and performed the experiments. JW, XT, LP and XW: data analysis. JW and YZ: wrote and revised the manuscript. All authors have accepted responsibility for the entire content of this manuscript and approved submission.

